# Polygenic associations with phenotypic classes across the psychosis-affective spectrum

**DOI:** 10.64898/2026.07.10.26357470

**Authors:** Charlotte A Dennison, Sophie E Legge, Alastair G Cardno, Diego Quattrone, Peter Holmans, Arianna Di Florio, Katherine Gordon-Smith, Ian Jones, Lisa Jones, Michael J Owen, Michael C O’Donovan, James TR Walters

## Abstract

**Introduction:** Limitations of current classifications of schizophrenia, schizoaffective disorder, and bipolar disorder are evident from their overlapping symptoms, aetiologies, treatments, and outcomes, and present a barrier to novel treatment discovery. Alternative conceptualisations are needed to address nosological validity, align diagnosis to aetiology, and improve prognostication and treatment choice. We aimed to identify latent classes across the psychosis spectrum based on premorbid functioning and outcomes, and assess these in relation to genetic liability and symptom dimensions.

**Method:** Participants with a diagnosis of schizophrenia, schizoaffective disorder, or bipolar disorder type 1, were ascertained from four UK clinical cohorts (total n=5,043). Latent class analysis was conducted using phenotypes not included within the diagnostic criteria, including premorbid functioning, age at illness onset, and measures of severity and course. Polygenic scores (PGS) for psychiatric disorders and behavioural traits were tested for associations with latent classes. We tested if diagnosis explained associations between PGS and classes.

**Results:** A three-class model provided the best fit. Class one had poorer premorbid functioning, lower rates of recovery, and higher PGS for schizophrenia and ADHD. Class three had the highest functioning, higher rates of psychosocial stressors before onset, higher intelligence PGS and lower PGS for psychiatric disorders. Class two was intermediate between classes one and three on measures of functioning, but was characterised by high levels of involuntary hospital admissions and high bipolar disorder PGS. Diagnosis only partially explained associations between PGS and class membership.

**Conclusions:** We identified classes across the psychosis spectrum characterised by different premorbid functioning and outcomes, that cut across diagnostic categories and captured genetic liability not explained by diagnosis. Our findings suggest alternative conceptualisations of psychotic disorders may complement diagnoses in mapping to the aetiology of these conditions, and could be useful to advance precision psychiatry.

## Introduction

Schizophrenia, schizoaffective disorder, and bipolar disorder substantially overlap in aetiology, risk factors, clinical presentation, and treatment^1^. As many as two thirds of individuals with bipolar disorder report experiencing psychosis during a mood episode^2^, whilst affective symptoms, particularly depression, are common in people with schizophrenia^3^. Schizoaffective disorder is defined by the presence of core symptoms of both schizophrenia and affective disorders, with psychotic symptoms also persisting for a period in the absence of mood symptoms^4^. Family studies have demonstrated familial aggregation of schizophrenia, schizoaffective disorder, and bipolar disorder^5^, and schizophrenia and bipolar disorder have a high genetic correlation based on commonly occurring risk variants(r_g_= 0.7)^6^, suggesting a large component of the genetic aetiology of these disorders is shared and that schizoaffective disorder represents an intermediate form on a schizophrenia-bipolar continuum^5,7^.

Within the psychosis-spectrum and indeed psychiatry in general, diagnostic classification is based on clinical signs and symptoms rather than underlying aetiology and pathophysiology. To improve prediction, and to advance efforts to identify new treatments and to target the most effective treatments to the right groups of individuals, novel methods of classifying people with these disorders that map more closely to the underlying biology are needed. Some studies have applied data-driven methods seeking to identify novel clusters across psychotic and affective disorders. These typically identify a severe subgroup characterised by high positive and negative symptoms with low levels of affective symptoms^8^. Others also report factors such as lower educational attainment and lower general functioning in this severe subgroup^9,10^. Recent work associated cognitive- and premorbid adjustment-based clusters with differing levels of environmental risk exposure, suggesting that socio-environmental factors may further contribute to heterogeneity across psychosis-spectrum presentations^11^. Other research has focused on clustering via cognitive and physiological data to identify ‘biotypes’ of psychosis^12^. Whilst assignment to these biotypes can be predicted using clinical data in a machine learning environment^13,14^, many of the measures defining the biotypes require complex assessments and are not readily available in all (or even most) healthcare settings. Other clusters identified by cross-disorder research vary substantially across studies, possibly due to variation in the measurements used, diagnoses included, and clustering methods employed. Few studies have tested for cluster replicability or sought to validate clusters using orthogonal data types, including genetic liabilities^15^.

Here, using a large sample of people with schizophrenia, schizoaffective disorder, and bipolar disorder, ascertained primarily from clinical services in the UK, we aimed to i) identify and validate latent classes of individuals across the psychosis spectrum based on premorbid functioning and outcome variables, ii) assess the relationship between classes and symptom dimensions, and iii) investigate the underlying biological validity of the classes by assessing the extent to which class membership is associated with polygenic scores (PGS) for neuropsychiatric disorders and intelligence.

## Methods

### Participants

Participants with schizophrenia, schizoaffective disorder, or bipolar disorder were ascertained from four cohorts: CardiffCOGS, Cardiff Affected-sibs, Cardiff F-series, and Bipolar Disorder Research Network (BDRN)^16,17^. Details on study recruitment are provided in the Supplementary Methods. All studies received relevant UK ethical approval, and all participants provided written informed consent.

### Phenotype data

Clinical assessment and data collection were closely aligned, by design, across the four cohorts. All participants were assessed using the Schedules for Clinical Assessment in Neuropsychiatry (SCAN)^18^, administered by a trained interviewer, and asked about other relevant clinical and demographic data. Trained researchers used information from the interview and, where available, secondary mental health records, to produce lifetime Operational Criteria Checklist (OPCRIT)^19^ ratings and derive lifetime research diagnoses based on ICD-10 criteria^4^. Individuals who met criteria for ICD-10 diagnoses of schizophrenia, schizoaffective disorder depressive type (SA-D), schizoaffective disorder bipolar type (SA-BP), and bipolar disorder type I were included in the present study. All studies showed good inter-rater reliability (kappa range 0.76-0.99)^17^.

### Genotype data

Individuals from CardiffCOGS, Cardiff Affected-sibs, and Cardiff F-series were genotyped using the Illumina HumanOmniExpress v8/v12 arrays. Individuals from BDRN were genotyped across three platforms: Illumina HumanOmniExpressExome-8, Affymetrix Genome-Wide Human SNP 6.0, Illumina PsychChip_v1.1. Following the initial quality control and imputation, described in the Supplementary Methods, samples were merged using PLINK v1.9, including only SNPs that were available across all three genotyping arrays with an INFO score >0.9 (n=3,398,557). A final round of quality control was applied to the merged data, excluding SNPs with: genotyping rate <0.98, Hardy-Weinberg equilibrium p-value <1×10^-6^, minor allele frequency <0.01, and sample missingness >0.05.

Genetic ancestry was identified using principal components analysis and a homogenous subgroup was retained, as described in Supplementary Methods. Identity by descent was conducted in PLINK v1.9 to identify related pairs with a kinship score greater than 0.15. One individual from each related pair was retained for analysis, prioritised by highest genotyping rate following QC.

### Polygenic scores

PGS were calculated using PRS-CS^20^ with scores based on the results from GWAS for schizophrenia, bipolar disorder, major depressive disorder (MDD), ADHD, autism, and intelligence, for further details see Supplementary Method. PGS were standardised against a control sample (Wellcome Trust Case Control Consortium^21^) with a mean of zero and standard deviation of one, in order to facilitate comparison across PGS.

### Patient and public involvement

People with lived experience who are members of the National Centre for Mental Health’s Partnership in Research (PÂR) group were consulted and helped formulate the research questions for these analyses. These ideas were further developed with people with lived experience and representative third sector organisations (Bipolar UK and Adferiad) as part of the Brain and Genomics Hub of the UKRI Mental Health Platform^22^.

## Analysis

### Latent class analysis

The R package ‘depmixS4’ was used to conduct latent class analysis (LCA), using the phenotypes in Table S1 as indicator variables. Phenotypes were selected for LCA on the basis that they were of clinical relevance, such as treatment response, or an indicator of functioning, such as premorbid social functioning. Importantly, to avoid deriving classes that simply mapped onto diagnostic labels, we did not include symptoms or any measures used to assign an ICD diagnosis. Phenotypes were required to be available and measured consistently across all four study cohorts.

The sample was randomly split into discovery and validation samples using the R package ‘randomizr’, with 70% of the sample assigned to the discovery group. Four latent class models, labelled A-D, based on different covariates were tested to explore their impact on class assignment and model fit. Definitions of all indicator variables and covariates are in Table S1. All models were run separately in the discovery and validation sample. Model A included all indicator variables in Table S1 with no covariates; model B included all indicator variables with age at interview as a covariate; model C included all indicator variables, with age at interview and source of rating as covariates; model D included all indicator variables, with age at interview, source of rating, and duration of illness as covariates. A two-class and a three-class solution was tested for each model; a four-class solution was also tested but this could not be identified for any model, so a higher number of class solutions was not tested. The ‘mix’ function was used to define each model, and the ‘multistart’ function to fit the model, using 50 starts and a maximum of 100 iterations of the expectation-maximisation algorithm per start. All models were tested only on participants with complete data for all three covariates used in model D to ensure tests of model fit, which are sensitive to sample size, were based on the same number of individuals. LCA tolerates missing data in the indicator variables, thus individuals were included regardless of whether they had complete or incomplete data for the indicator variables. Model fit was compared using the Bayesian Information Criterion (BIC), which has been shown to be the most reliable indicator of model fit^23^.

### Confirmatory factor analysis

To allow us to investigate the relationships between latent classes and symptoms, confirmatory factor analysis (CFA) of symptom data was performed in R using the ‘lavaan’ package. OPCRIT data from all samples were used to create a five-factor model, with factors representing positive, negative, disorganised, manic, and depressive symptoms, described in the Supplementary Methods. We chose a five-factor model as these factors are often found in factor analysis of psychotic and affective symptoms^24^.

### Genetic associations

Logistic regression was used to analyse the relationships between class membership and PGS for schizophrenia, bipolar disorder, MDD, ADHD, autism, and intelligence in the full sample. Models were run for each PGS separately, and as a multivariable model including all PGS.

To investigate whether associations between class and PGS could be explained by diagnosis, we used linear regression to measure the association between PGS and class membership, covarying for diagnosis. We used linear regression with PGS as the outcome, as opposed to logistic regression with class as the outcome as in the previous models, in order to jointly measure the effect of class and diagnosis.

To ensure that PGS associations with symptom dimensions and latent classes were not due to batch effects resulting from the use of different genotyping arrays, PGS analyses in the full sample were repeated using only individuals genotyped using the Illumina Omni-Express array (n=2,957). For all PGS analyses, we included the first five PCs as covariates, as well as any of the first 20 PCs that were associated (p<0.05) with the outcome.

## Results

Of 5,043 individuals included in the study (54% female, mean age 45.5 years), 2,975 (59%) had a main ICD-10 research diagnosis of bipolar affective disorder (type 1), 1,557 (31%) had a diagnosis of schizophrenia, 317 (6%) had a diagnosis of SA-BP, and 194 (4%) had a diagnosis of SA-D. Of these, 4,114 (81.6%) were included in the latent class analysis as they had complete data for the three covariates. The demographics of each cohort are presented in Table S3.

### Latent class analysis

For both the discovery and validation samples, the best fit based on the BIC value (Table S4) was a three-class solution of Model D. Individuals were assigned to an optimum class where their probability of membership of that class was >50%. Those who could not be so assigned were excluded from analysis (n=48, 1.2%). Distribution of class membership probability is displayed in Figure S2. A comparison of the classes between the discovery and validation samples indicated that the models were identifying highly similar latent classes, and thus the discovery and validation samples were combined for subsequent analysis. Figures S3 and S4 display the distribution of each phenotype across the classes, stratified by sample.

Within each of the latent classes, the proportions of individuals exhibiting the categorical phenotypes entered into the LCA are displayed in Figure 1. Means and frequencies of all phenotypes included in the LCA are displayed in Figures S3 and S4. Almost all variables showed a gradient between classes, with class two intermediate between classes one and three, except for ‘ever admitted under the mental health act (MHA)’, which was highest in class two, and alcohol dependency, which showed less variation across the classes than the other variables.

**Figure 1.**
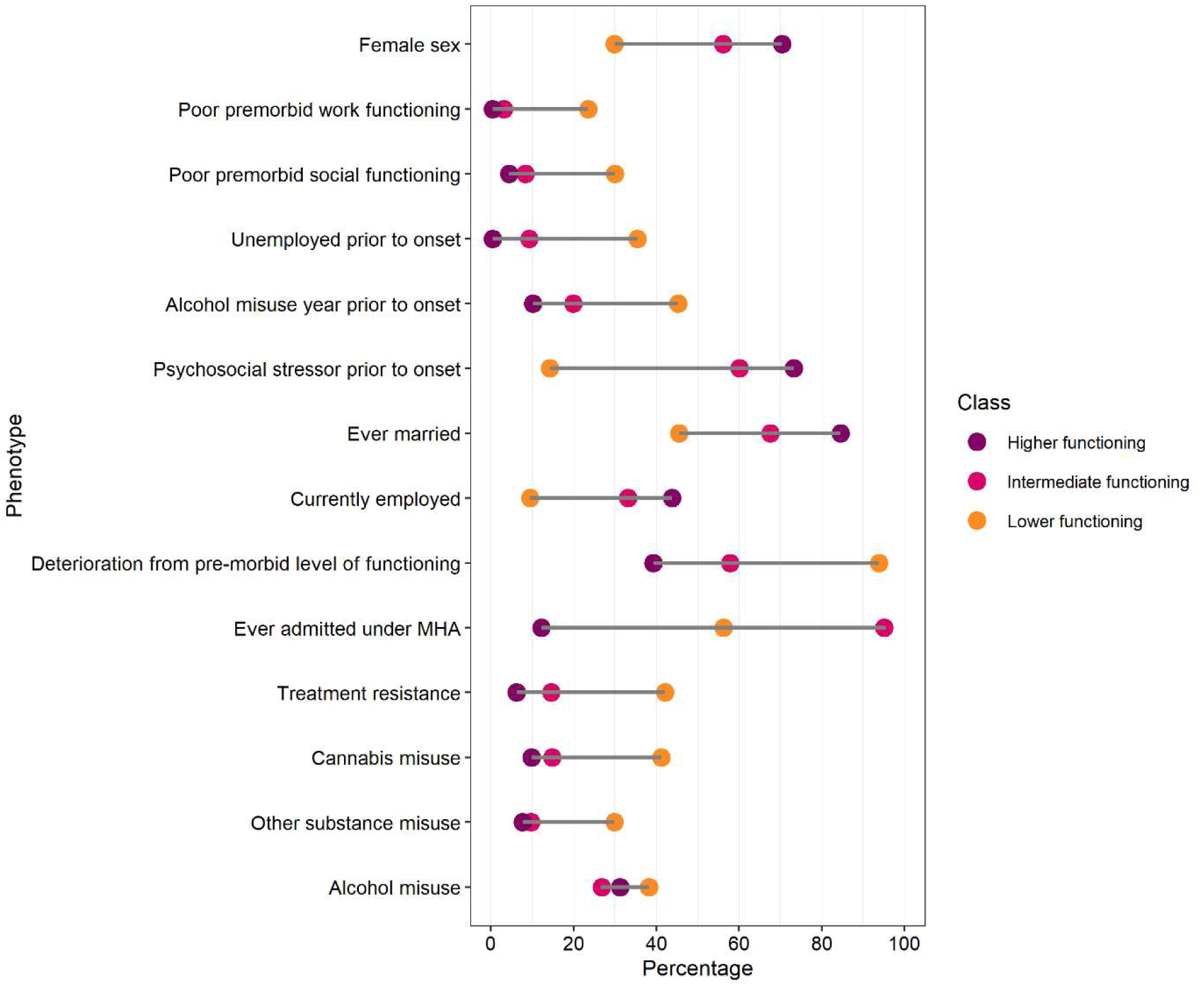
Percentage of individuals in each class with the listed phenotypes. Purple dots refer to the higher functioning class, pink dots refer to the intermediate functioning class, and orange dots refer to the lower functioning class.

Class 1 (n=1166) had the highest occurrence of items indexing poorer premorbid functioning, including lower educational attainment, and had a high proportion of males. Class 1 was further characterised by poorer outcomes than the other two classes, with the lowest rates of current employment, highest rates of treatment resistance, and almost all individuals experiencing deterioration from their premorbid level of functioning. Given the pattern observed, we refer to class 1 as the ‘lower functioning’ class.

Class 3 (n=1393), had the highest rates of items indicating better premorbid functioning than the other classes, including the highest rates of employment prior to onset, and had a high proportion of females. This group also had the highest rate of individuals who reported a psychosocial stressor in the six months prior to onset, and the lowest rates of deterioration from premorbid level of functioning. Given the observed pattern, we refer to class 3 as the ‘higher functioning’ class.

We refer to class 2 (n=1507) as the ‘intermediate functioning’ class as, by most metrics, it was intermediate between classes 1 and 3 with the exception that it had the highest rate of compulsory admission, with almost all individuals having been admitted under the Mental Health Act.

Table 1 describes the numbers and frequencies of ICD diagnoses in each latent class. Most, but by no means all, people with schizophrenia and SA-D were in the lower functioning class, whilst individuals with bipolar disorder were relatively evenly divided between the intermediate and higher functioning classes (Table 1). Individuals with SA-BP were more evenly split across the classes.

**Table 1.** Number of individuals with each diagnosis in each class. Percentage refers to the proportion of individuals with that diagnosis assigned to each class.

| **Diagnosis** | **Lower**<br>**functioning class** | **Intermediate**<br>**functioning class** | **Higher**<br>**functioning class** | **TOTAL** |
| --- | --- | --- | --- | --- |
| Schizophrenia | 837 (64%) | 379 (29%) | 90 (7%) | **1306** |
| SA-D | 97 (55%) | 59 (34%) | 20 (11%) | **176** |
| SA-BP | 93 (36%) | 93 (36%) | 71 (28%) | **257** |
| Bipolar disorder | 139 (6%) | 976 (42%) | 1212 (52%) | **2327** |
| **TOTAL** | **1166** | **1507** | **1393** |  |

### Symptom dimensions and latent classes

Results of the CFA are summarised in Tables S2 and S4, and Figure S5. Mapping symptom dimensions onto the latent classes (Figure 2), revealed that the lower functioning class had higher rates of positive, negative, and disorganised symptoms, with lower rates of depressive and manic symptoms. Conversely, the higher functioning class had lower rates of positive, negative, and disorganised symptoms, and higher rates of depressive and manic symptoms. The intermediate class had moderate levels of each symptom dimension (Figure 2).

**Figure 2.**
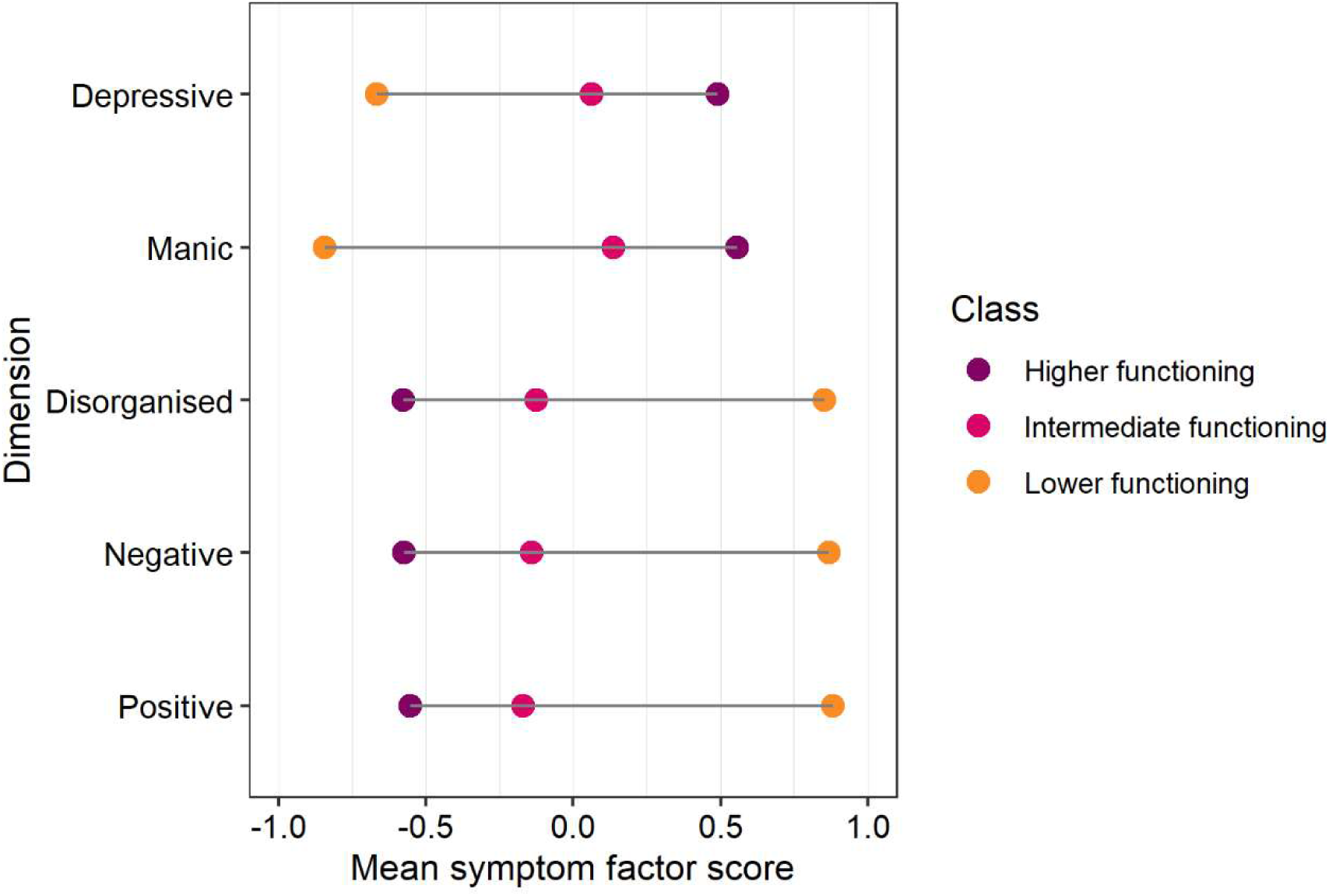
Mean standardised symptom factor score by class. Purple dots refer to means in the higher functioning class, pink dots refer to means in the intermediate functioning class, and orange dots refer to means in the lower functioning class.

### PGS associations with class membership

High quality genotype data were available for 3,131 (low functioning class n=698; intermediate functioning class n=1189; higher functioning class n=1244) unrelated participants of European genetic ancestry who were successfully assigned an optimum class. The mean PGS for each disorder/trait per class is displayed in Figure 3, for reference the mean PGS per diagnosis is displayed in Figure S6. Results of class membership regressed on PGS are displayed in Table 2. Lower schizophrenia PGS and higher intelligence PGS were associated with better levels of functioning. Pairwise comparisons revealed that schizophrenia PGS was different between all class pairs, and intelligence PGS was different between the higher functioning class and both the lower and intermediate classes. MDD and ADHD PGS were higher in the lower functioning class compared with each of the other two classes (Table 2), but were not significantly different between the intermediate and higher functioning classes. In contrast, bipolar disorder PGS showed a complex pattern, being highest in the intermediate group, which differed from both lower and higher functioning classes, despite the higher functioning class containing a greater proportion (52%) of people with bipolar disorder than the intermediate class (42%).

**Figure 3.**
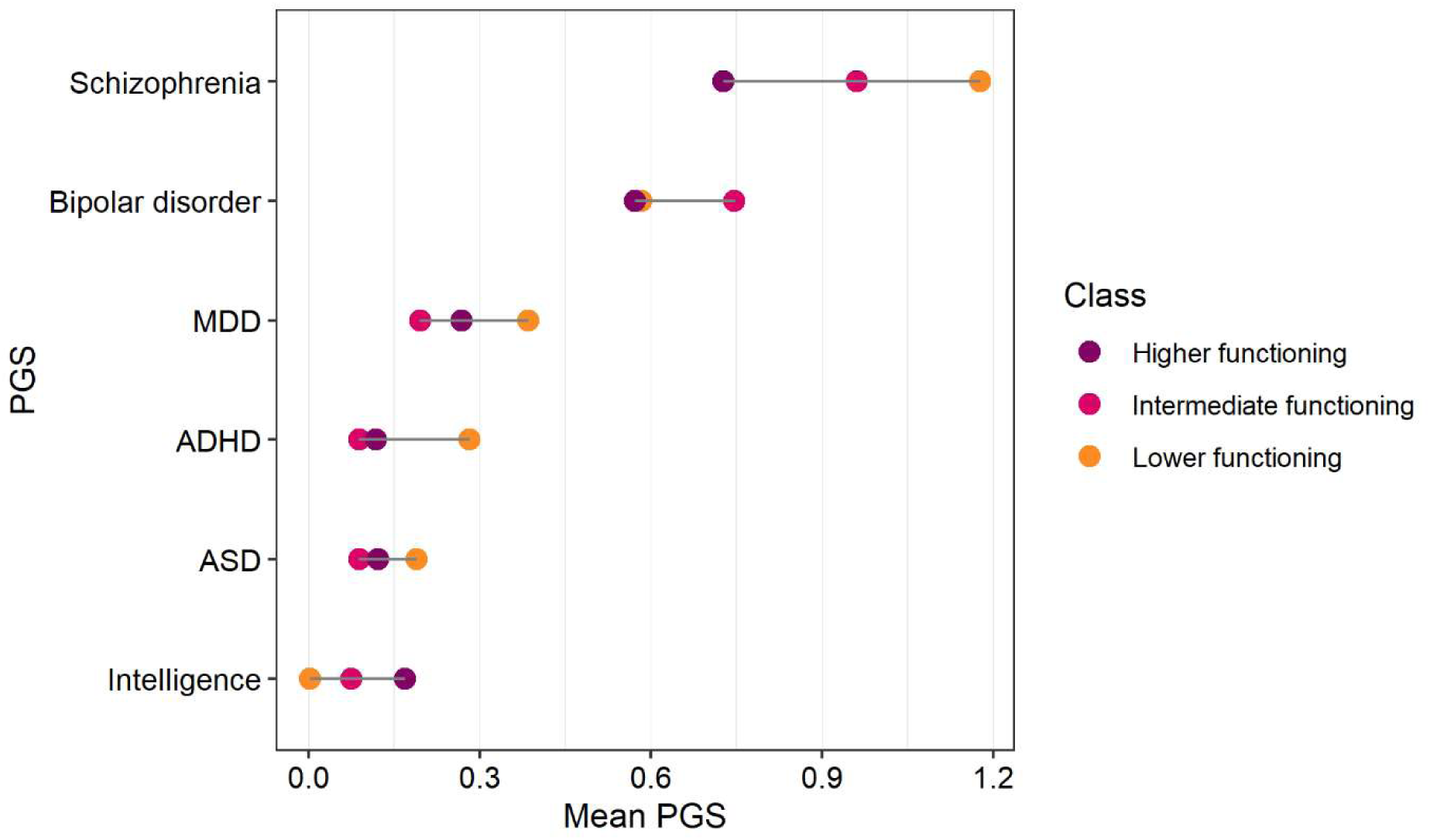
Mean standardised PGS for each disorder/trait by class. Purple dots refer to means in the higher functioning class, pink dots refer to means in the intermediate functioning class, and orange dots refer to means in the lower functioning class.

**Table 2.** Association between PGS and latent class membership using univariable models. Odds ratio refers to the first class named, with the second class as the reference category.

| Class Comparison | PGS | OR | Lower CI | Upper CI | P-value |
| --- | --- | --- | --- | --- | --- |
| Lower vs intermediate functioning | Schizophrenia | 1.20 | 1.09 | 1.32 | 2.9E-04 |
|  | Bipolar disorder | 0.84 | 0.76 | 0.92 | 3.8E-04 |
|  | MDD | 1.17 | 1.07 | 1.29 | 1.1E-03 |
|  | ADHD | 1.17 | 1.06 | 1.29 | 1.4E-03 |
|  | Autism | 1.10 | 1.00 | 1.21 | 0.062 |
|  | Intelligence | 0.97 | 0.88 | 1.06 | 0.47 |
| Lower vs higher functioning | Schizophrenia | 1.53 | 1.39 | 1.70 | 9.8E-17 |
|  | Bipolar disorder | 0.99 | 0.90 | 1.08 | 0.77 |
|  | MDD | 1.10 | 1.00 | 1.21 | 0.060 |
|  | ADHD | 1.13 | 1.03 | 1.24 | 0.014 |
|  | Autism | 1.04 | 0.95 | 1.15 | 0.41 |
|  | Intelligence | 0.88 | 0.80 | 0.97 | 0.008 |
| Intermediate vs higher functioning | Schizophrenia | 1.26 | 1.16 | 1.37 | 4.9E-08 |
|  | Bipolar disorder | 1.17 | 1.08 | 1.26 | 1.3E-04 |
|  | MDD | 0.93 | 0.86 | 1.01 | 0.073 |
|  | ADHD | 0.96 | 0.88 | 1.04 | 0.30 |
|  | Autism | 0.97 | 0.89 | 1.05 | 0.45 |
|  | Intelligence | 0.92 | 0.85 | 1.00 | 0.040 |

To investigate the effect of correlation between PGS on the results, we conducted multivariable analyses of PGS, displayed in Table S6. The effect sizes were mostly consistent with the univariable models, although some findings were slightly stronger, such as between lower bipolar disorder PGS and the lower functioning vs other two classes.

To examine whether the latent classes were able to capture variation in PGS in addition to that captured by diagnosis, we regressed both class membership and diagnosis on PGS. Full results are displayed in Table S7. When covarying for diagnosis, the lower functioning class continued to be associated with elevated ADHD PGS relative to both classes, whilst the higher functioning class continued to be associated with lower bipolar disorder PGS relative to both the other classes. The higher functioning class also had lower schizophrenia PGS and higher MDD PGS than the intermediate functioning class.

All effect sizes from the analysis restricted to samples genotyped only using the Illumina Omni-Express array were consistent with the primary estimate, indicating that the results of the latter were not driven by unadjusted array effects (Table S6). Although p-values did increase as a result of the reduced sample size.

## Discussion

Using independent discovery and validation samples of individuals with schizophrenia, schizoaffective disorder, and bipolar disorder, we identified three classes characterised by distinct patterns of premorbid functioning, outcomes, and polygenic liability. Classes were defined by shared phenotypes that are not used as criteria to define diagnosis, and each class contained a mixture of individuals with each diagnosis, suggesting that subgroups may exist that are partly independent of existing categorical diagnoses.

The lower functioning class had poorer premorbid functioning and worse outcomes, including treatment resistance and deterioration from premorbid level of functioning. Previous studies have identified a subgroup of individuals characterised by more severe psychotic and affective symptoms and lower educational attainment^9,10^. Our finding of a lower functioning class associated with higher psychotic symptoms and lower affective symptoms supports these studies and further suggests that this group may have a stronger neurodevelopmental aetiology, given the higher ADHD and schizophrenia PGS, relative to the other classes. It is possible that this class is comparable to ‘biotype 1’ identified by the B-SNIP consortium^13,25^, which was associated with high levels of positive, negative, and disorganised symptoms and lower levels of depressive symptoms^26^. However, differences in the measures used to define classes across our study and B-SNIP preclude direct comparison. The lower functioning class also had lower bipolar disorder PGS alongside higher schizophrenia PGS, relative to the other classes. This is notable given that schizophrenia and bipolar disorder have a strong positive genetic correlation^6^, thus a disparity between these PGS (or a PGS that differentiates schizophrenia from bipolar disorder^27^) could potentially be a useful marker of poorer outcomes.

The higher functioning class was characterised by better outcomes, including higher rates of employment and recovery to premorbid level of functioning, as well as higher intelligence PGS and lower schizophrenia PGS than the other two classes. The higher functioning class is most consistent with previous observations of clusters termed ‘classic mania’ or ‘pure mania’, characterised by higher mood symptoms alongside better functioning and outcomes than is typically seen in mania with psychosis^28,29^. Notably, the higher functioning class had the highest rates of a psychosocial stressor in the six months prior to onset, which could suggest that individuals in this class are more responsive to stress, relative to individuals in the other classes, and that exposure to stress could be an important route into psychosis for this group. Schizophrenia PGS and psychotic symptoms were lowest in this class, consistent with observations that schizophrenia PGS correlates with psychotic symptoms across the schizophrenia-bipolar spectrum^7,27^.

The intermediate class had levels of functioning that fell between the other two classes, and was associated with intermediate schizophrenia PGS and high bipolar disorder PGS relative to the other classes. Moreover, despite intermediate levels of symptomatology (Figure 2), this group had a remarkably high rates of involuntary hospitalisation. One possible explanation for this observation is that, whilst this group did not experience the highest levels of any one symptom domain, the combination of appreciable levels of both psychotic and affective symptoms could be more likely to result in an involuntary admission, compared to higher levels of psychotic or affective symptoms alone. However, involuntary admission rates have increased over time and vary by age, sex, ethnicity, and post-code^30^, and it is possible that different ascertainment strategies across the cohorts could have influenced this result. Thus, this finding requires replication in external datasets.

We note that individuals from each diagnostic group are distributed across classes suggesting we are capturing information beyond that provided by clinical diagnosis alone. We demonstrated that PGS differentially associate across classes and that this is not wholly explained by diagnosis. In this respect our classes seem to be identifying groups of individuals with genetic architectures partly independent of diagnoses and supports further genetic investigation of such classes.

### Strengths and limitations

Our sample sizes across diagnostic groups are a major strength, and in contrast to previous research we were able to include individuals with schizoaffective disorder, distinguishing between depressive and bipolar subtypes. A further strength is the fact that the studies were conducted in the same centre by overlapping teams, and that the protocols, data collection and ratings procedures were aligned across studies and that common instruments (e.g. OPCRIT) were used. However, we were unable to include some potentially important phenotypes due to lack of data availability across samples. In particular, cognitive functioning could be a useful stratifier between classes, but this was not available in most datasets. Whilst we performed the latent class analysis in separate discovery and validation samples derived from our four cohorts, our study is limited by the lack of an independent validation sample and thus we cannot assess the transferability of our findings across settings.

The cohorts included in our study recruited predominantly people of European genetic ancestry, and further research in other ancestral groups is essential to replicate and expand our findings, as the distribution of stratifiers, symptoms, and PGS may differ across populations.

### Conclusion

Our findings suggest that clusters of individuals characterised by phenotypes such as similar premorbid functioning and outcomes can be identified across the psychosis-affective spectrum that are partly independent of diagnosis. Whilst diagnosis explains a large degree of the association between genetic liability and class, the classes are able to capture phenotypic homogeneity across diagnoses that is associated with genetic liability not fully captured by current diagnostic systems.

## Acknowledgements and Funding

This work was supported by funding from the National Institute of Mental Health (1U01MH109514-01).

This work was supported by a UKRI MRC Programme Grant (MR/Y004094/1) to JTRW, SEL, PH, MJO and MCOD and the UKRI Mental Health Platform Brain and Genomics Hub grant (MR/Z503745/1) to JTRW, SEL and PH.

CD is funded by the Wolfson Centre for Young People’s Mental Health, established with support from the Wolfson Foundation.

BDRN funding was supported by grants from the Wellcome Trust [078901] and the Stanley Medical Research Institute [6045240-5500000100].

## Declaration of interests

JTRW, MJO and MCOD have received grant funding from Takeda Pharmaceuticals and Akrivia Health for work unrelated to the current research.

## Data availability statement

The data used in this study is not publicly available due to ethical restrictions. If researchers wish to discuss access, please contact the authors.

## Author contributions

CD – conceptualisation, formal analysis, methodology, writing – original draft

SL – conceptualisation, funding acquisition, methodology, supervision, writing – review & editing AC – investigation, methodology, writing – review & editing

DQ – methodology, software, writing – review & editing

PH - funding acquisition, methodology, writing – review & editing

ADF - methodology, writing – review & editing

KGS – data curation, methodology, writing – review & editing

IJ - methodology, writing – review & editing

LJ - methodology, writing – review & editing

MO – funding acquisition, methodology, writing – review & editing

MOD – conceptualisation, funding acquisition, methodology, supervision, writing – review & editing

JW – conceptualisation, funding acquisition, methodology, supervision, writing – review & editing

## Notes

### Author Declarations

The South East Wales Research Ethics Committee Panel and the UK National Health Service Research Authority gave ethical approval for this work.

