## Supplementary Methods for "Polygenic associations with phenotypic classes across the psychosis-affective spectrum"

**Supplementary material**

### **Supplementary methods**

#### *Participant recruitment*

Individuals with a clinical diagnosis of schizophrenia or a schizophrenia-spectrum disorder were recruited to CardiffCOGS^1^ from 2009 - 2017 and Cardiff F-series^2^ from 1997 - 2001 via secondary mental health services. The Cardiff Affected-sibs study^3^ recruited families from 1994 - 1997 with two or more siblings. One sibling was required to have a diagnosis of schizophrenia or schizoaffective disorder depressive-type (SA-D), but we additionally included sibships where the other member had schizoaffective disorder bipolar-type (SA-BP). BDRN recruited individuals with a clinical bipolar-spectrum diagnosis from secondary mental health services, lithium clinics, and patient support organisations from 2001–2023. For this study, only individuals with genotype data were included, which were those participants who had been recruited between 2001-2013^4^.

#### *Initial genotype data quality control and imputation*

We ascertained a sample of control participants from the Wellcome Trust Case Control Consortium^5^, for the purpose of creating a reference sample from which to standardise polygenic scores. Control participants were not screened for psychiatric disorders, however it has previously been demonstrated that if as many as 5% of the sample were to have or develop any of the disorders of interest, that this would constitute a loss of power comparable to reducing the sample size by around 10%^1^. Given that schizophrenia and bipolar disorder type 1 each have a lifetime prevalence of <1%^2,3^, for case-control analyses, greater power is achieved by using a large unscreened set of controls than using a typically much smaller set of screened controls. Controls were genotyped using the Affymetrix GeneChip 500K Mapping Array Set through the Wellcome Trust Case-Control Consortium (WTCCC)^1^.

In the first stage, samples underwent quality control (QC) checks prior to imputation. Genotype Harmoniser v1.42 was used to align SNPs against the Haplotype Reference Consortium (HRC) panel v1.1. SNPs with discordant information were updated to match the reference panel, in order to maximise the number of SNPs available for imputation. Sex checks were performed in PLINK; discordant results were manually checked to determine whether the sex was inaccurately recorded or whether sample contamination had occurred. Where this could not be determined, the sample was excluded from further analysis. Finally, the following exclusion filters were applied: SNP call rate <0.95, participant genotyping rate <0.95, SNPs with Hardy-Weinberg Equilibrium (HWE) p-value<1x10^-6^, SNPs with minor allele frequency (MAF) <0.01.

Samples genotyped on the same array were imputed together, regardless of original study. Imputation was conducted on the Michigan Imputation Server using the HRC v1.1 reference panel^4^. Following imputation, a second round of SNP QC filters were applied, SNPs were excluded if: genotype probability <0.9 per individual, MAF <0.01, genotyping rate <0.95, HWE p-value<1x10^-4^, and INFO score <0.3.

#### *Principal components analysis and identity by descent*

SNPs were pruned for identity by descent (IBD) and principal components analysis (PCA), retaining one SNP from any pair of SNPs with an R^2^ >0.2 in a 500kb window. PCA was performed in PLINK v1.9 to investigate the effects of multiple genotyping platforms and to select individuals of European genetic ancestry. The function ‘covMCD’ from the R package ‘robustbase’^6^ was used to identify individuals with European ancestry, using the first six principal components (PCs). Individuals with a distance within the 95^th^ percentile of the centre point of the multi-dimensional space were retained for polygenic analyses (281 cases excluded). Figure S1 summarises the results of the PCA, including PCs before and after selecting for ancestry. We did not identify effects related to genotyping platform.

#### *Polygenic scores*

PRS-CS auto^7^ was used to derive polygenic scores, using summary statistics from GWAS for schizophrenia^8^, bipolar disorder^9^, major depressive disorder (MDD)^10^, ADHD^10^, autism^11^, and intelligence^12^. As samples used in this study were also included in the GWAS for schizophrenia and bipolar disorder, custom sets of summary statistics for these phenotypes were derived by the Psychiatric Genomics Consortium Working Groups that excluded these samples.

#### *Confirmatory factor analysis*

In the latent class analysis, we excluded items that typically inform a diagnosis of schizophrenia, schizoaffective disorder, or bipolar disorder to allow us to investigate whether other phenotypes could identify clusters independent of diagnosis. Thus, we excluded diagnostic symptoms, family history of psychiatric illness, and number of episodes of psychosis, mania, and depression. Subsequently, to investigate how the latent classes related to symptoms, we conducted confirmatory factor analysis (CFA) using OPCRIT^13^ data from all samples to create a five-factor model. The five factors represented positive symptoms, negative symptoms, disorganised symptoms, mania, and depression. We used confirmatory, as opposed to exploratory factor analysis, as several studies have previously applied factor analysis to symptom data in psychotic and affective disorders, and these factors are widely regarded to represent the spectrum of symptoms in schizophrenia and bipolar disorder^14–18^. We assigned items to their specific factor based on previous literature and prior theoretical knowledge^15,16^.

CFA model selection was undertaken in two stages - model selection and fitting in R, and implementation of final model in MPlus. First, we used the ‘cfa’ function in the R package ‘lavaan’^19^ to create a five-factor model. We set the parameters of the model such that factors were allowed to be correlated and were standardised to have a mean of zero and a variance of one^17^. All other parameters remained in the default setting for lavaan^19^. OPCRIT items rated positively in over 5% of individuals and missing in fewer than 20% were included in the initial model (Supplementary Table 2). For subsequent iterations of the model, items were removed on the basis of high loading onto multiple factors (indicating that an item is associated with more than one factor), or high correlation between two items, in which instance the item with the lowest missingness was retained. This process was repeated until adjustments suggested by the modification indices did not substantially improve the model fit. Model fit was compared using several fit indices: i) Comparative Fit Index (CFI), ii) Tucker Lewis Index (TLI), iii) Root Mean Square Error of Approximation (RMSEA), and iv) Standardised Root Mean square Residual (SRMR). The CFI measures how well the model fits the data compared to a null model. The TLI is similar in that it compares to a baseline model, but differs in that the TLI penalises an overly complex model. For both the CFI and TLI, higher values indicate better fit, with a value >0.9 considered good fit^20^. RMSEA measures how closely the model fit compares to a perfect fitting model, by comparing the deviation of the chi-square statistic from the degrees of freedom. Lower RMSEA values indicate closer proximity to a perfect model, and values <0.05 indicate a good fit^20^. SRMR is similar to RMSEA except that it is a standardised measure that does not consider the degrees of freedom when comparing the observed and expected covariance matrices. Lower SRMR values indicate better fit, with values <0.05 indicating good fit^21^. The best fitting model, based on model fit indices, was retained for subsequent analysis. Only individuals with complete data can be included in the lavaan model, meaning that factors scores were available for 2,163 individuals. Therefore, in the second stage, we implemented the best fitting model in MPlus using the maximum likelihood estimator and Montecarlo integration to obtain factor scores for the full sample. The first four models (Table S2) were not tested in MPlus as Montecarlo integration on five factors is computationally demanding. We correlated factor scores between the lavaan and MPlus methods to assess whether factor scores generated by MPlus were similar to those generated by lavaan.

### **Supplementary Tables**

#### **Table S1.** Definition of each item included in the latent class analysis.

Includes total number and percentage of individuals with data for the item.

| Variable | Description | N (5043) |
| --- | --- | --- |
| Age first contact | Age at which first contact was made with secondary psychiatric services. | 3773 (75%) |
| Educational attainment | Highest educational attainment: 0 = none, 1 = 11+, 2 = CSE, 3 = O-Level or GCSE, 4 = A-level, 5 = Degree | 4544 (90%) |
| Suicidal ideation | Rated most severe lifetime-ever. 0 = absent, 1 = tedium vitae, 2 = suicidal ideation, 3 = suicide attempt unlikely to result in death  4 = suicide attempt likely to result in death  5 = multiple suicide attempts likely to result in death | 3851 (76%) |
| Lowest Global Assessment Scale (GAS) | Lifetime worst GAS score in a psychotic, manic, or depressive episode. Higher score indicates better functioning. | 3235 (64%) |
| Highest occupation | Lifetime highest occupation.  1 = Unemployed/unable to work, 2= manual/trade work, 3= professional.  Groupings are based on ONS Standard Occupational Classification (Professional = classes 1-4, manual/trade = classes 5-10). | 3099 (61%) |
| Current occupation | Occupation at interview.  1 = Unemployed/unable to work, 2= manual/trade work, 3= professional. | 3043 (60%) |
| Sex | Self-reported sex. 1 = male, 2 = female. | 5033 (99%) |
| Married | Ever been married. 0 = no, 1= yes. | 4409 (97%) |
| Ever detained under the mental health act | Ever detained under section 2 or 3 of the Mental Health Act. | 3850 (76%) |
| Alcohol abuse within year prior to onset | Alcohol abuse where quantity is excessive, where alcohol related complications occur, during the year prior to first psychiatric contact. OPCRIT item 12. | 4279 (85%) |
| Unemployed at onset | Unemployed at onset of illness. Parent working full time in the home scored as employed. Students attending classes on full time course, scored as employed. OPCRIT item 7. | 4138 (82%) |
| Poor premorbid work adjustment | Prior to illness onset, unable to keep any job for more than 6 months, had a history of frequent changes of job or was only able to sustain a job well below that expected by educational level or training. OPCRIT item 9. | 4198 (83%) |
| Poor premorbid social adjustment | Difficulty entering or maintaining normal social relationships, showed persistent social isolation, withdrawal or maintained solitary interests prior to onset of illness. OPCRIT item 10 | 4056 (80%) |
| Psychosocial stressor prior to onset | A severely or moderately severely threatening event has occurred prior to onset of disorder that is unlikely to have resulted from the individual’s own behaviour (i.e., the event can be seen as independent or uncontrollable). Stressor must have been experienced within 6 months prior to onset. 0=no, 1=yes. OPCRIT item 16. | 3586 (71%) |
| Cannabis misuse | One of the following must have occurred persistently for at least one month: continued use despite knowledge of having a persistent or recurrent social, occupational, psychological or physical problem that is caused or exacerbated by cannabis; or recurrent use in situations in which it is physically hazardous; or symptoms definitely indicative of dependence. OPCRIT item 82. | 4128 (82%) |
| Substance misuse | One of the following must have occurred persistently for at least one month: Continued use despite knowledge of having a persistent or recurrent social, occupational, psychological or physical problem that is caused or exacerbated by substance use; or recurrent use in situations in which it is physically hazardous; or symptoms definitely indicative of dependence. Relates specifically to any substance other than cannabis or alcohol, which are covered by other items. OPCRIT item 83. | 4180 (83%) |
| Deterioration from premorbid functioning | Individual does not regain their premorbid social, occupational, or emotional functioning after an acute episode of illness. OPCRIT item 88. | 3983 (79%) |
| Treatment resistance | Evidence of resistance to either antipsychotic or lithium therapy. For antipsychotic resistance, 0= substantial improvement in psychotic symptoms either subjectively or according to medical records, or if relapse occurs when medication is stopped. Rated 1 if person did not meet these criteria or was treated with Clozapine for treatment resistance. OPCRIT item 89.  Lithium resistance defined as no subjective or objective evidence of beneficial response to lithium treatment.  0 = responsive, 1 = resistant. | 4114 (82%) |
| Alcohol misuse | One of the following must have occurred persistently for at least one month: Continued use despite knowledge of having a persistent or recurrent social, occupational, psychological or physical problem that is caused or exacerbated by alcohol; or recurrent use in situations in which it is physically hazardous; or symptoms definitely indicative of dependence. OPCRIT item 81. | 3964 (79%) |
| Age at interview | Age at interview in years. Used as a covariate. | 4932 (98%) |
| Source of rating | 1= Hospital case notes (charts).  2= Structured interview with subject [rated only no case-notes have been obtained]  3= Prepared abstract  4= Interview with informant  5= Combined sources including structured interview  6= Combined sources not including structured interview  OPCRIT item 1.  Used as a covariate. | 4492 (89%)  By source:  1 = 1%  2 = 22%  3 = <1%  4 = <1%  5 = 71%  6 = 6% |
| Duration of illness | Duration of time since illness onset. Calculated as age at interview - age at first contact with secondary services.  Used as a covariate. | 3721 (73%) |

#### **Table S2.** OPCRIT items included in each CFA model.

Columns indicate OPCRIT item, corresponding factor, total number and percentage with data, percentage positively endorsing the item, and whether the item was included in each model.

| Factor | OPCRIT item | N (%) | % Positive | Included in Model 1 | Included in Model 2 | Included in Model 3 | Included in Model 4 | Included in Model 5 |
| --- | --- | --- | --- | --- | --- | --- | --- | --- |
| Depression | 50. Increased appetite | 4323 (85.7%) | 26.6 | Yes | No | No | No | No |
|  | 23. Agitated activity | 4248 (84.2%) | 36.0 | Yes | Yes | Yes | No | No |
|  | 37. Dysphoria | 4834 (95.9%) | 85.2 | Yes | Yes | No | No | No |
|  | 46. Early wakening | 4265 (84.6%) | 36.5 | Yes | No | No | No | No |
|  | 47. Excessive sleep | 4256 (84.4%) | 40.9 | Yes | No | No | No | No |
|  | 44. Initial insomnia | 4390 (87.1%) | 51.1 | Yes | No | No | No | No |
|  | 25. Loss of energy | 4561 (90.4%) | 73.6 | Yes | Yes | Yes | Yes | Yes |
|  | 39. Loss of pleasure | 4594 (91.1%) | 75.7 | Yes | Yes | Yes | Yes | Yes |
|  | 45. Middle insomnia | 4274 (84.8%) | 44.8 | Yes | No | No | No | No |
|  | 48. Poor appetite | 4486 (89.0%) | 56.5 | Yes | No | No | No | No |
|  | 41. Poor concentration | 4496 (89.2%) | 73.7 | Yes | Yes | Yes | Yes | Yes |
|  | 42. Self-reproach | 4494 (89.1%) | 68.0 | Yes | No | Yes | Yes | Yes |
|  | 24. Slowed activity | 4187 (83.0%) | 46.2 | Yes | Yes | Yes | Yes | Yes |
|  | 43. Suicidal ideation | 4716 (93.5%) | 74.2 | Yes | Yes | Yes | Yes | No |
|  | 51. Weight gain | 4193 (83.1%) | 22.8 | Yes | No | No | No | No |
|  | 49. Weight loss | 4249 (84.3%) | 38.8 | Yes | No | No | No | No |
| Disorganised | 17. Bizarre behaviour | 4407 (87.4%) | 37.6 | Yes | No | No | No | No |
|  | 28. Positive formal thought disorder | 4695 (93.1%) | 14.0 | Yes | Yes | Yes | Yes | Yes |
|  | 34. Inappropriate affect | 4816 (95.5%) | 9.3 | Yes | Yes | Yes | Yes | Yes |
|  | 26. Speech difficult to understand | 4696 (93.1%) | 25.0 | Yes | Yes | Yes | No | No |
| Mania | 21. Distractibility | 4594 (91.1%) | 59.1 | Yes | Yes | Yes | Yes | Yes |
|  | 35. Elation | 4842 (96.0%) | 69.4 | Yes | Yes | Yes | Yes | Yes |
|  | 19. Excess activity | 4805 (95.3%) | 66.7 | Yes | Yes | Yes | Yes | Yes |
|  | 53. Increased sociability | 4521 (89.6%) | 55.4 | Yes | Yes | Yes | Yes | Yes |
|  | 36. Irritable | 4741 (94.0%) | 58.8 | Yes | Yes | Yes | Yes | Yes |
|  | 30. Pressured speech | 4818 (95.5%) | 67.6 | Yes | Yes | Yes | Yes | Yes |
|  | 20. Reckless activity | 4499 (89.2%) | 55.1 | Yes | Yes | Yes | Yes | Yes |
|  | 22. Reduced need for sleep | 4790 (95.0%) | 65.7 | Yes | Yes | Yes | Yes | Yes |
|  | 56. Self-esteem | 4626 (91.7%) | 61.5 | Yes | Yes | Yes | Yes | Yes |
|  | 31. Thoughts racing | 4797 (95.1%) | 68.0 | Yes | Yes | Yes | Yes | Yes |
| Negative | 29. Negative formal thought disorder | 4812 (95.4%) | 11.5 | Yes | Yes | Yes | Yes | Yes |
|  | 32. Restricted affect | 4811 (95.4%) | 18.0 | Yes | Yes | Yes | Yes | Yes |
| Positive | 73. 3^rd^ person auditory hallucinations | 4542 (90.1%) | 18.8 | Yes | Yes | Yes | Yes | Yes |
|  | 59. Bizarre delusions | 4787 (94.9%) | 13.5 | Yes | Yes | No | No | No |
|  | 58. Delusions of influence | 4356 (86.4%) | 62.2 | Yes | Yes | No | No | No |
|  | 76. Nonaffective auditory hallucinations | 4311 (85.5%) | 32.1 | Yes | No | No | No | No |
|  | 54.Persecutory delusions | 4469 (88.6%) | 47.8 | Yes | Yes | Yes |  | Yes |
|  | 75. Persecutory voices | 4528 (89.8%) | 31.7 | Yes | Yes | Yes | Yes | Yes |
|  | 74. Running commentary | 4581 (90.8%) | 8.1 | Yes | No | No | No | No |
|  | 67. Thought withdrawal | 4849 (96.2%) | 5.2 | Yes | Yes | No | No | No |
|  | 61. Passivity | 4721 (93.6%) | 8.9 | Yes | Yes | Yes | Yes | Yes |
|  | 63. Primary delusions | 4830 (95.8%) | 11.1 | Yes | No | No | No | No |
|  | 66. Thought insertion | 4721 (93.6%) | 10.8 | Yes | Yes | Yes | Yes | Yes |

#### **Table S3.** Demographic information on each cohort included in the study.

|  | | **Cardiff COGS** | **Cardiff Affected-Sibs** | **Cardiff F-series** | **Bipolar Disorder Research Network** |
| --- | --- | --- | --- | --- | --- |
| **Total sample size** | | 1001 | 381 | 636 | 3025 |
| **Female sex** | | 374 (37.4%) | 127 (33.3%) | 198 (31.1%) | 2018 (66.7%) |
| **Mean age (SD)** | | 43.3 (11.9) | 40.9 (12.4) | 41.7 (14.1) | 47.5 (12.2) |
| **Main lifetime ICD-10 diagnosis** | **Schizophrenia** | 713 (71.2%) | 330 (86.6%) | 514 (80.8%) | 0 (0%) |
|  | **SA-D** | 151 (15.1%) | 24 (6.3%) | 19 (3.0%) | 0 (0%) |
|  | **SA-BP** | 95 (9.5%) | 23 (6.0%) | 36 (5.7%) | 163 (5.4%) |
|  | **Bipolar disorder** | 42 (4.2%) | 4 (1.0%) | 67 (10.5%) | 2862 (94.6%) |

#### **Table S4.** Bayesian Information Criterion (BIC) values for each latent class model tested in the discovery and validation samples.

A smaller number indicates a better fit.

| **Model** | **Number of classes** | **Discovery**  **BIC** | **Validation**  **BIC** |
| --- | --- | --- | --- |
| A | 2 | 89725 | 38300 |
|  | 3 | 38943 | 15852 |
| B | 2 | 89567 | 38260 |
|  | 3 | 34275 | 16113 |
| C | 2 | 89515 | 38227 |
|  | 3 | 36470 | 16018 |
| D | 2 | 89486 | 38226 |
|  | 3 | 33347 | 15642 |

#### **Table S5.** Fit indices for all CFA models tested using lavaan.

Columns refer to Comparative Fit Index (CFI), Tucker-Lewis Index (TLI), Root Mean Square Error of Approximation (RMSEA), Standardised Root Mean Square Residual (SRMR). For CFI and TLI, values closer to one indicate better fit, with a value >0.9 considered indicative of good fit. For RMSEA and SRMR, lower values indicate better fit, with values <0.05 considered a good fit. All models are five-factor models with varying items included, specific items included in each model are specified in Table S2.

| **Model** | **CFI** | **TLI** | **RMSEA (95% CI)** | **SRMR** |
| --- | --- | --- | --- | --- |
| One | 0.996 | 0.996 | 0.077 (0.075 - 0.078) | 0.113 |
| Two | 0.999 | 0.999 | 0.032 (0.030 - 0.035) | 0.057 |
| Three | 1 | 1 | 0.024 (0.022 - 0.027) | 0.044 |
| Four | 1 | 1 | 0.017 (0.013 - 0.020) | 0.033 |
| Five | 1 | 1 | 0.015 (0.012 - 0.019) | 0.032 |

#### **Table S6.** Association between PGS and latent class membership.

Displayed for the primary univariable analysis, multivariable analysis, and when restricted to samples genotyped using the Omni-Express array (univariable, n=2,500). Odds ratio refers to the first classed named, with the second class as the reference category.

| **Class Comparison** | **PGS** | **Primary univariable analysis** | | | | **Multivariable models** | | | | **Omni sample only** | | | |
| --- | --- | --- | --- | --- | --- | --- | --- | --- | --- | --- | --- | --- | --- |
|  |  | **OR** | **Lower CI** | **Upper CI** | **P-value** | **OR** | **Lower CI** | **Upper CI** | **P-Value** | **OR** | **Lower CI** | **Upper CI** | **P-Value** |
| Lower vs intermediate functioning | Schizophrenia | 1.20 | 1.09 | 1.32 | 2.9E-04 | 1.28 | 1.15 | 1.42 | 3.9E-06 | 1.16 | 1.05 | 1.29 | 0.01 |
|  | Bipolar disorder | 0.84 | 0.76 | 0.92 | 3.8E-04 | 0.75 | 0.67 | 0.83 | 3.8E-08 | 0.85 | 0.76 | 0.94 | 2.0E-03 |
|  | MDD | 1.17 | 1.07 | 1.29 | 1.1E-03 | 1.13 | 1.01 | 1.25 | 0.03 | 1.18 | 1.06 | 1.31 | 2.1E-03 |
|  | ADHD | 1.17 | 1.06 | 1.29 | 1.4E-03 | 1.14 | 1.02 | 1.27 | 0.02 | 1.16 | 1.04 | 1.28 | 0.01 |
|  | Autism | 1.10 | 1.00 | 1.21 | 0.06 | 1.05 | 0.95 | 1.17 | 0.32 | 1.12 | 1.01 | 1.24 | 0.04 |
|  | Intelligence | 0.97 | 0.88 | 1.06 | 0.47 | 1.01 | 0.92 | 1.12 | 0.78 | 0.95 | 0.86 | 1.05 | 0.34 |
| Lower vs higher functioning | Schizophrenia | 1.53 | 1.39 | 1.70 | 9.8E-17 | 1.61 | 1.44 | 1.80 | 1.1E-17 | 1.55 | 1.40 | 1.73 | 1.1E-15 |
|  | Bipolar disorder | 0.99 | 0.90 | 1.08 | 0.77 | 0.82 | 0.74 | 0.91 | 2.6E-04 | 0.99 | 0.90 | 1.10 | 0.87 |
|  | MDD | 1.10 | 1.00 | 1.21 | 0.06 | 1.03 | 0.92 | 1.14 | 0.65 | 1.05 | 0.95 | 1.17 | 0.33 |
|  | ADHD | 1.13 | 1.03 | 1.24 | 0.01 | 1.07 | 0.96 | 1.20 | 0.20 | 1.11 | 1.00 | 1.22 | 0.05 |
|  | Autism | 1.04 | 0.95 | 1.15 | 0.41 | 1.02 | 0.91 | 1.13 | 0.77 | 1.05 | 0.95 | 1.17 | 0.33 |
|  | Intelligence | 0.88 | 0.80 | 0.97 | 0.01 | 0.93 | 0.84 | 1.02 | 0.14 | 0.88 | 0.80 | 0.98 | 0.01 |
| Intermediate vs higher functioning | Schizophrenia | 1.26 | 1.16 | 1.37 | 4.9E-08 | 1.23 | 1.12 | 1.34 | 7.5E-06 | 1.35 | 1.22 | 1.49 | 2.5E-09 |
|  | Bipolar disorder | 1.17 | 1.08 | 1.26 | 1.3E-04 | 1.12 | 1.03 | 1.21 | 0.01 | 1.17 | 1.07 | 1.28 | 5.8E-04 |
|  | MDD | 0.93 | 0.86 | 1.01 | 0.07 | 0.89 | 0.82 | 0.97 | 0.01 | 0.90 | 0.82 | 0.98 | 0.02 |
|  | ADHD | 0.96 | 0.88 | 1.04 | 0.30 | 0.96 | 0.88 | 1.05 | 0.36 | 0.96 | 0.88 | 1.05 | 0.39 |
|  | Autism | 0.97 | 0.89 | 1.05 | 0.45 | 0.99 | 0.90 | 1.08 | 0.78 | 0.96 | 0.87 | 1.06 | 0.42 |
|  | Intelligence | 0.92 | 0.85 | 1.00 | 0.04 | 0.92 | 0.85 | 1.00 | 0.05 | 0.94 | 0.86 | 1.03 | 0.16 |

#### **Table S7.** Association between PGS and latent class membership when covarying for diagnosis.

Beta refers to the first classed named, with the second class as the reference category.

| **Class Comparison** | **PGS** | **Beta** | **Lower CI** | **Upper CI** | **P-value** |
| --- | --- | --- | --- | --- | --- |
| Lower vs intermediate functioning | Schizophrenia | -0.06 | -0.17 | 0.05 | 0.32 |
|  | Bipolar disorder | -0.01 | -0.12 | 0.11 | 0.89 |
|  | MDD | 0.11 | 0.00 | 0.22 | 0.055 |
|  | ADHD | 0.12 | 0.01 | 0.23 | 0.040 |
|  | Autism | 0.09 | -0.02 | 0.20 | 0.12 |
|  | Intelligence | -0.01 | -0.12 | 0.11 | 0.92 |
| Lower vs higher functioning | Schizophrenia | 0.12 | -0.02 | 0.26 | 0.093 |
|  | Bipolar disorder | 0.22 | 0.08 | 0.37 | 2.6E-03 |
|  | MDD | 0.13 | -0.01 | 0.27 | 0.076 |
|  | ADHD | 0.14 | 0.00 | 0.29 | 0.047 |
|  | Autism | -0.01 | -0.15 | 0.13 | 0.93 |
|  | Intelligence | -0.10 | -0.24 | 0.05 | 0.21 |
| Intermediate vs higher functioning | Schizophrenia | 0.15 | 0.07 | 0.23 | 2.2E-04 |
|  | Bipolar disorder | 0.21 | 0.12 | 0.29 | 1.2E-06 |
|  | MDD | -0.09 | -0.17 | 0.00 | 0.042 |
|  | ADHD | -0.04 | -0.13 | 0.04 | 0.29 |
|  | Autism | -0.03 | -0.11 | 0.05 | 0.47 |
|  | Intelligence | -0.08 | -0.16 | 0.00 | 0.065 |

### **Supplementary Figures**

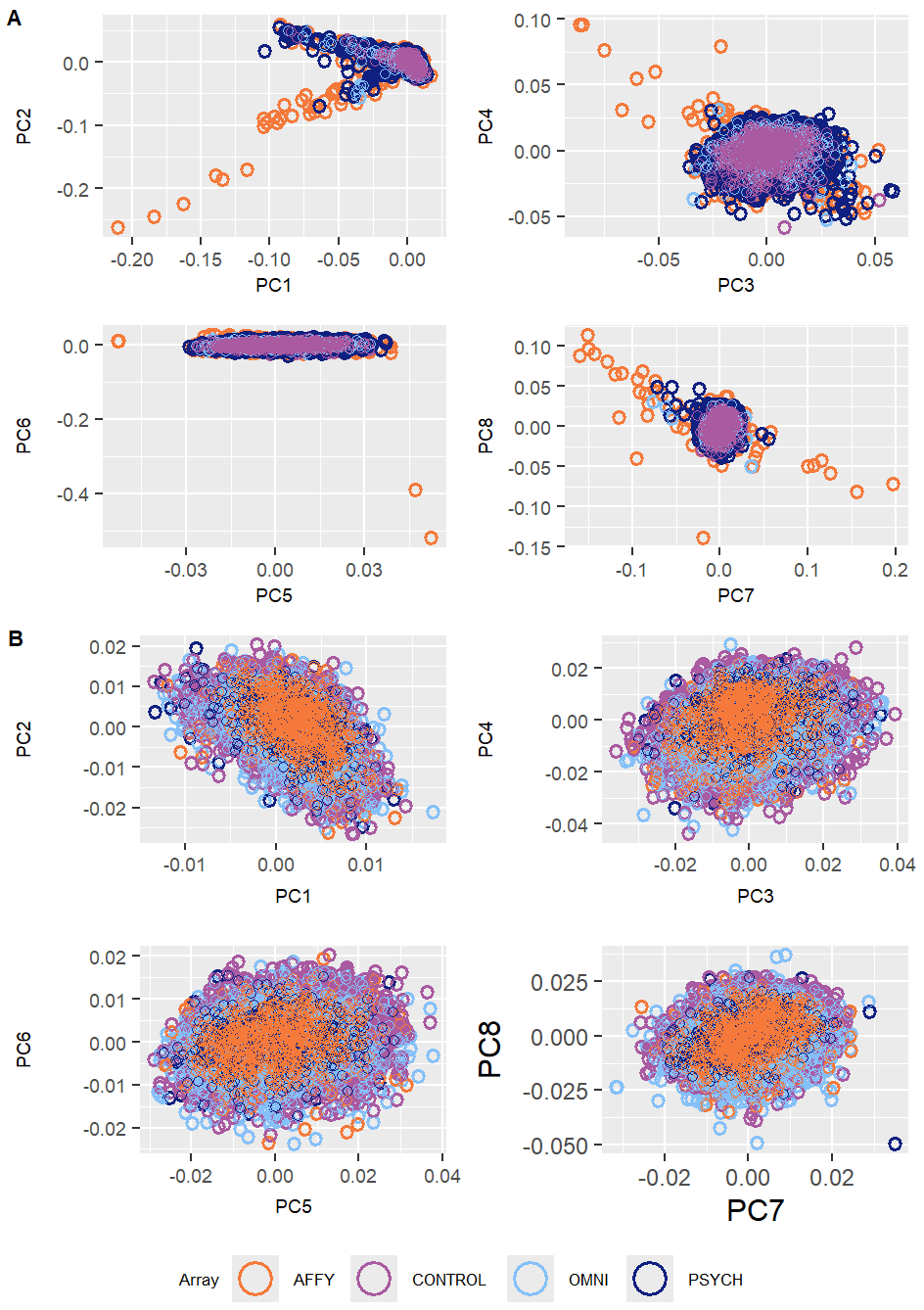

#### **Figure S1**. Principal components analysis

A) Principal components (PC) one to eight, colour indicates genotyping platform (red = Affymetrix, green=Omni-express, blue=Psych chip). B) Principal components (PC) one to eight after selecting for European ancestry, colour indicates genotyping platform.

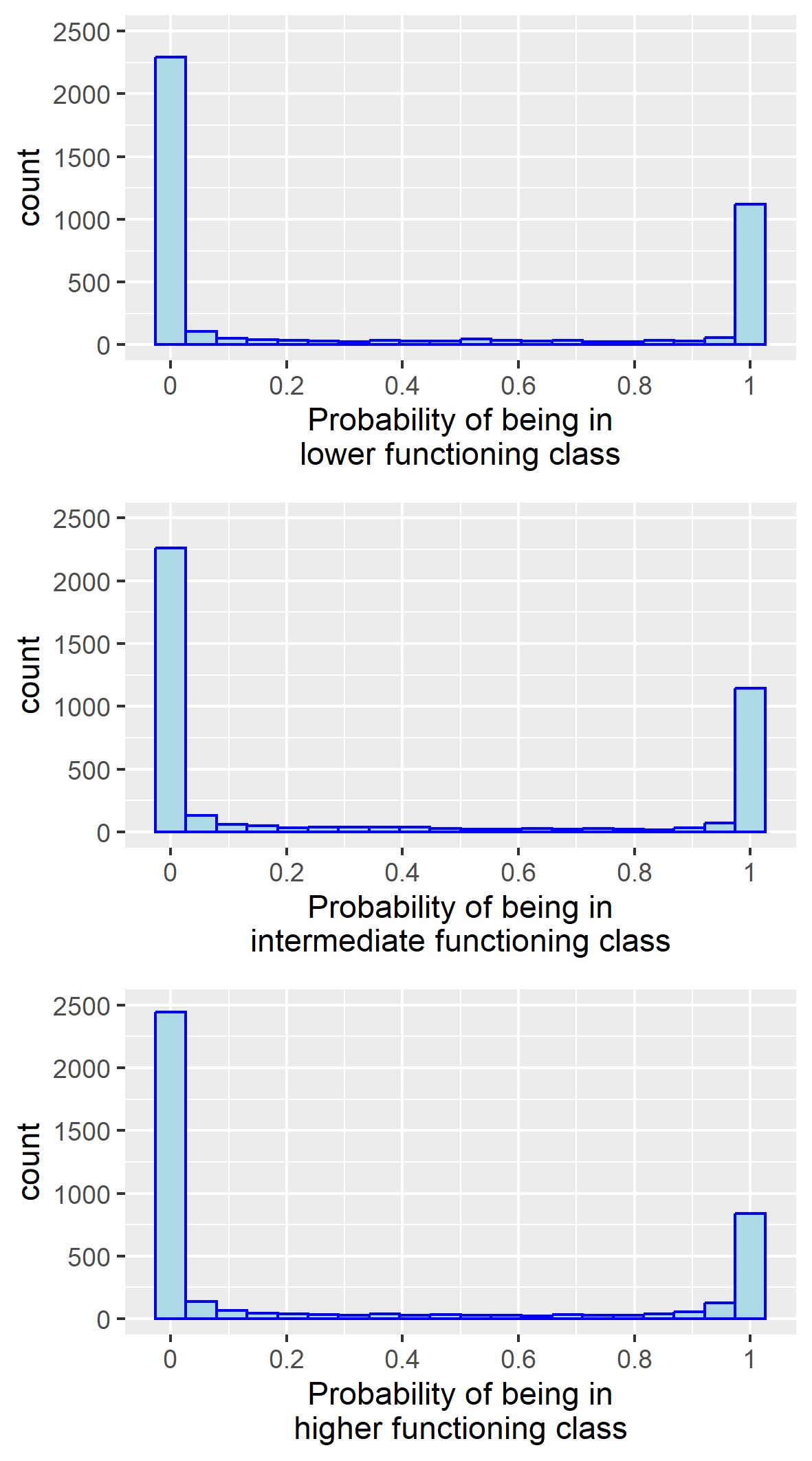

#### **Figure S2.** Distribution of the probability of being assigned to each latent class.

All individuals are assigned a probability of belonging to each of the three classes, and individuals were assigned to their ‘optimum class’ if they had a 50% or higher probability of belonging to that class. Individuals with <50% probability for every class were excluded (n=48).

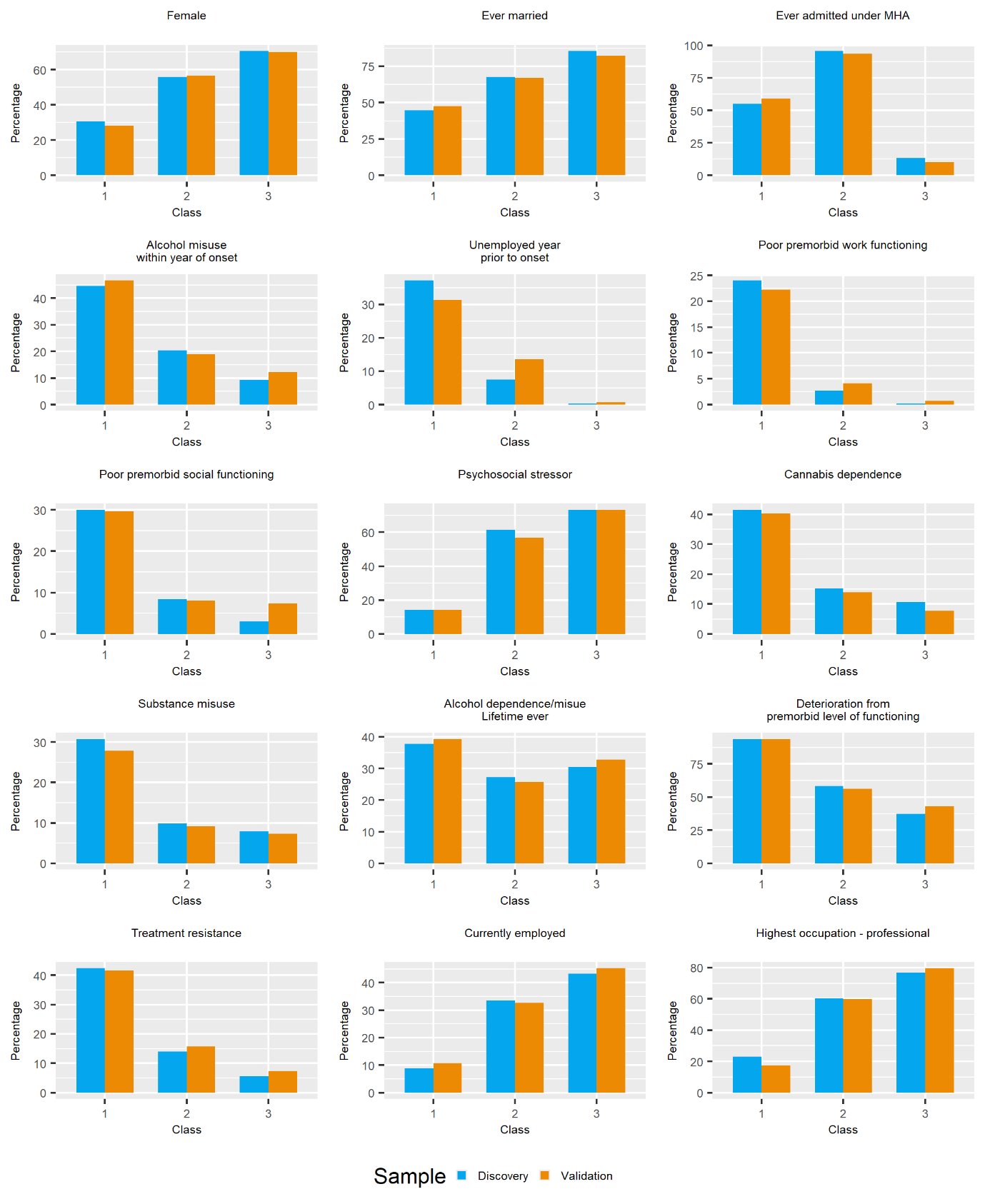

**Figure S3.** Percentage of individuals in each class, stratified by sample, endorsing each of the categorical phenotypes included in the latent class analysis.

Blue indicates the discovery sample, whilst orange shows the validation sample.

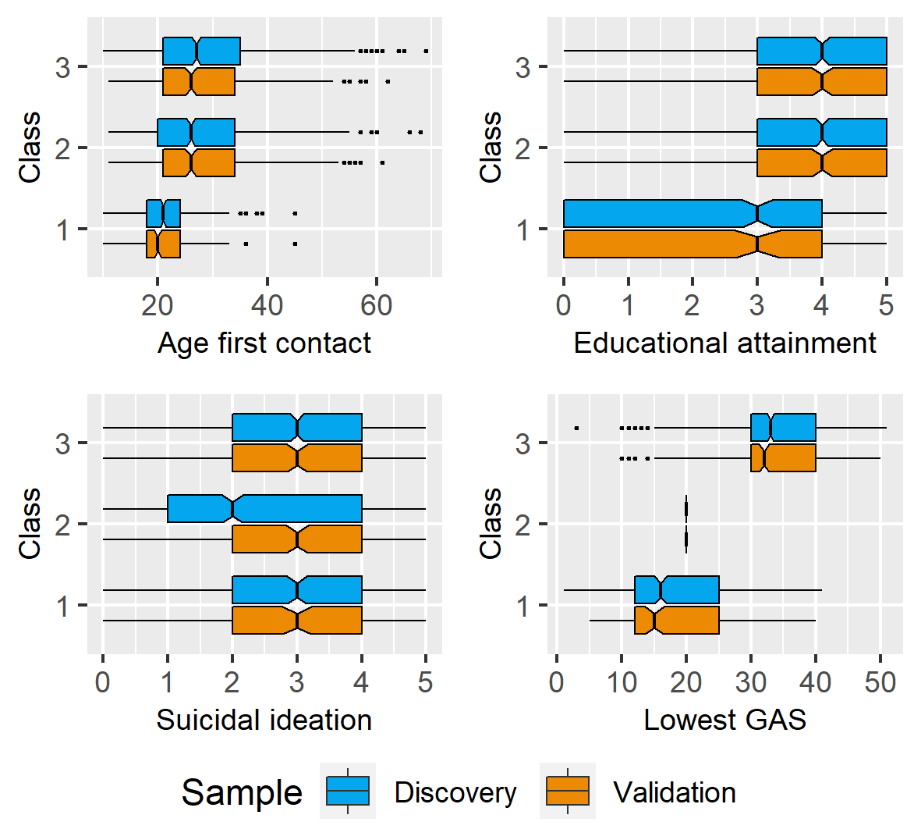

#### **Figure S4.** Boxplots of the median, interquartile range, minimum and maximum values, and outliers for the continuous items included in the model, by class.

Blue indicates distribution in the discovery sample, whilst orange shows the validation sample.

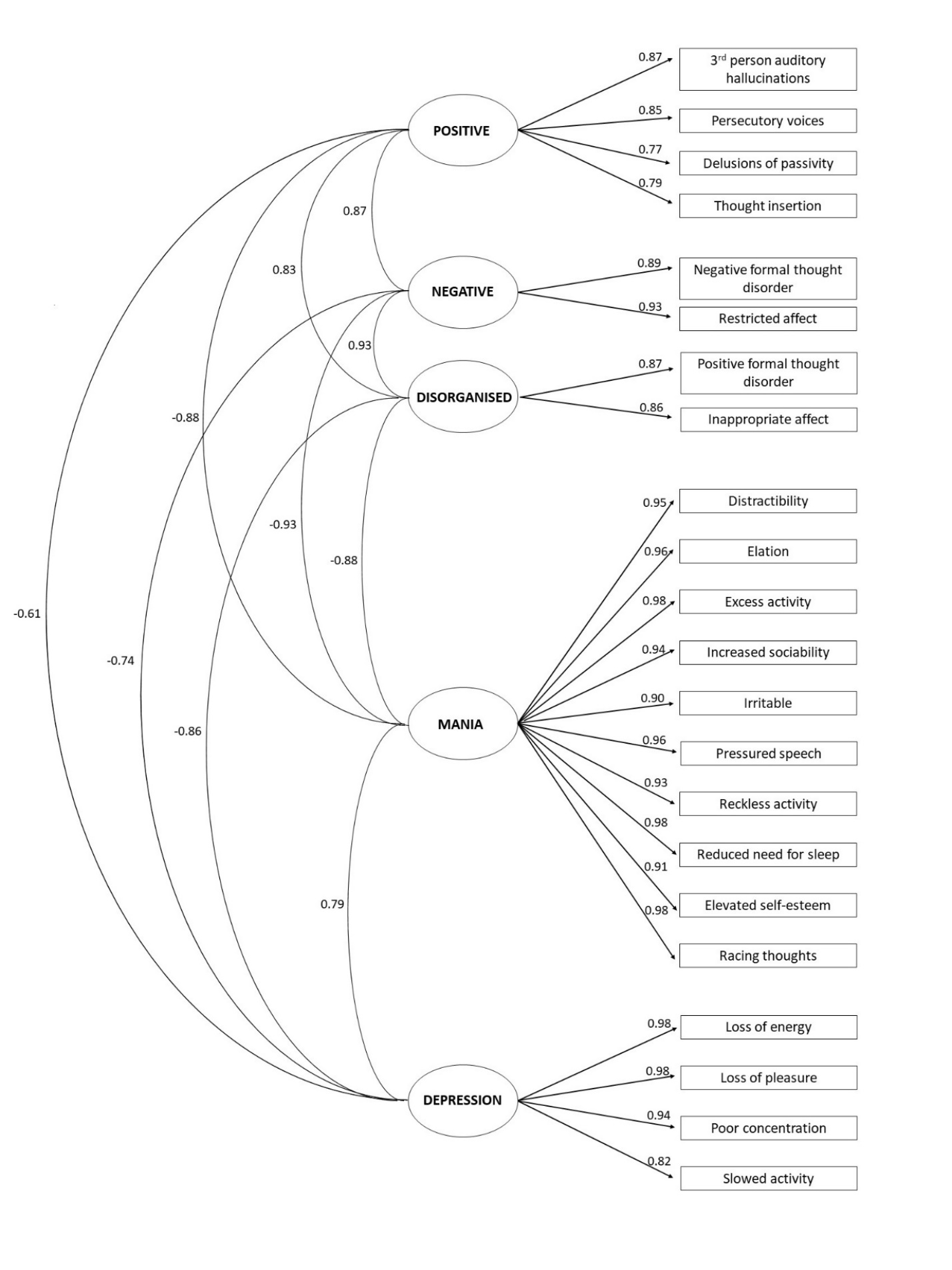

#### **Figure S5.** Structure of confirmatory factor analysis for model five, the best fitting model, in MPlus.

Values indicate the loading of each item onto the corresponding factor and correlations between factors.

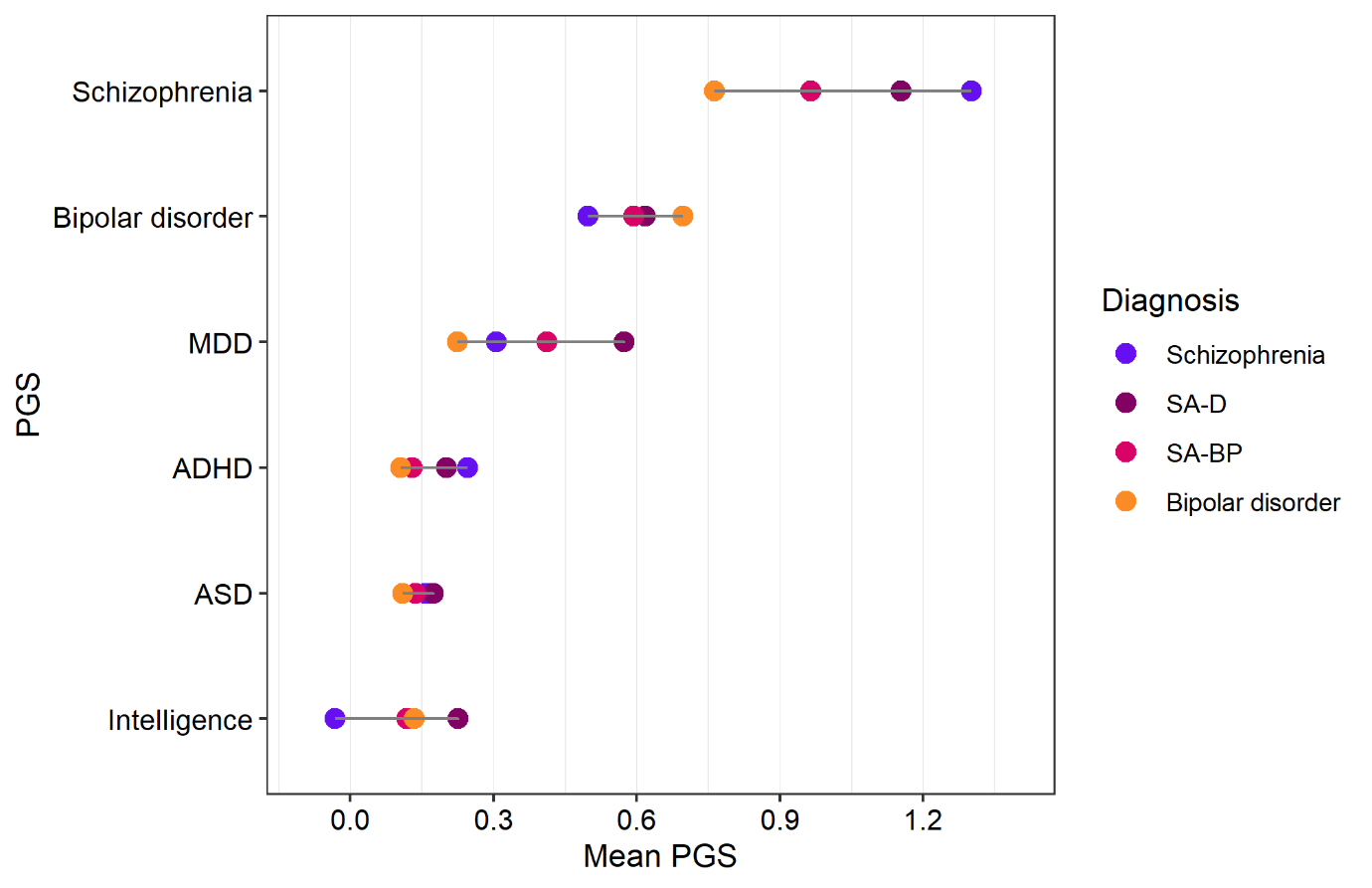

#### **Figure S6.** Mean polygenic score (PGS) for each disorder/trait by diagnosis.

Points indicate mean and colour indicates research diagnosis of schizophrenia (blue), schizoaffective disorder depressive type (SA-D, purple), schizoaffective disorder bipolar type (SA-BP, pink), and bipolar disorder (orange). PGS were standardised against a population mean of zero and standard deviation of one.
